# Performance of a rapid SARS-COV-2 serology test in whole blood and separated plasma

**DOI:** 10.1101/2020.10.02.20199083

**Authors:** S. Vemulapati

## Abstract

Rapid SARS-COV-2 related serology testing can help identify and manage the spread of infection in decentralized testing environments but the limitation in performance of existing tests in blood has restricted implementation of testing at the point-of-care. Optimization of existing rapid tests in whole blood will require significant effort in the short-term and there is a need for solutions to help bridge the gap in performance between plasma and whole blood. We demonstrate here the implementation of the H.E.R.M.E.S platform, a portable plasma separation system that can enhance the performance of blood-based diagnostic testing, with a commercially available SARS-COV-2 IgG/IgM serology rapid diagnostic test (RDT) in a blinded study with 61 human samples. We compare the performance of the RDT in whole blood and separated plasma and highlight that plasma yields a 39% increase in positivity agreement with PCR in samples collected from patients with early infections. We further legitimize the increase in positivity agreement rate with the help of an independent evaluation by 10 previously untrained users. The H.E.R.M.E.S plasma separation system circumvents the need for assay optimization in whole blood and furthers the legitimacy of incorporating SARS-COV-2 serology RDTs at the point-of-care. The data highlighted in this work makes a compelling case for the incorporation of the H.E.R.M.E.S system in large scale efforts to perform SARS-COV-2 serology testing in decentralized testing environments.

## Introduction

As of September 15th 2020, the novel coronavirus (SARS-COV-2) has infected over 28 million people worldwide^1^. Serology testing has emerged as an effective avenue to monitor the spread and prevalence of infection in communities and healthcare settings^2–5^ since IgG and IgM seroconversion can be reliably observed in individuals infected with SARS-CoV-2 within 2-17 days of initial symptom onset^6–10^. As opposed to molecular testing, serology testing can be used as an epidemiological tool to gather information on infection history across population clusters and help enhance contact tracing efforts in the event of a localized outbreak^11,12^. To expedite availability of diagnostic tests to the public, the Food and Drug Administration (FDA) issued an Emergency Use Authorization (EUA) on Feb 21 2020 allowing immunoassay and serology tests to be available commercially based on manufacturer-reported data without formal clearance and approval^13^. To date, the FDA has issued EUAs for 31 different serology tests^14^; a third of which are further classified as lateral flow rapid diagnostic tests (RDT) which offer the benefit of ease of operation, low sample volumes and quick results (10-15 minutes)^15–17^.

RDTs are the ideal platform to facilitate mass testing as they can be easily distributed and incorporated in decentralized testing environments. However, RDTs are currently limited in application for SARS-COV-2 serology testing at the point-of-care because of issues with their performance. While manufacturers claim equivalent diagnostic accuracy in blood, serum and plasma, the majority of self-reported validation data has been collected by testing serum or plasma samples alone. Several lab studies have shown that the actual performance of these tests is much lower than advertised^18–20^ which has called the legitimacy of serology testing into question^21^. Additionally, under the current EUA, there are no serology tests approved for use in point-of-care settings which would further suggest that existing tests have not demonstrated reliable performance in venous and fingerstick blood samples. RDTs are currently restricted for use in moderate and high complexity labs^22^, which are diametrically opposite in function to point-of-care testing environments and are likely have access to more robust lab-based tests. The limitation in performance has ultimately bottlenecked the application of RDTs for use at the point-of-care and a widescale rollout of RDT serology tests will require demonstration of robust and reliable performance in blood.

The difficulty to demonstrate clinically acceptable performance in blood stems from the interference introduced by red blood cells, which can make up to 55% of the total volume of the sample^23^. While the cell themselves adversely affect traditional immunochemical diagnostic methods^24,25^, red blood cell separation itself is a tedious process that is not compatible with facilities and resources typically available in point-of-care settings. To address the gap in available solutions for sample processing in decentralized testing environments, we previously demonstrated the High Efficiency Rapid Magnetic Erythrocyte Separator (H.E.R.M.E.S) portable plasma separation platform: a low-cost magnetic bead capture assay to remove up to 99.9% of all red blood cells in a sample to deliver plasma that can enhance the diagnostic accuracy of lateral flow tests that are designed for use with blood^26^. In this work, we demonstrate the implementation of the H.E.R.M.E.S platform with a commercially available FDA EUA approved SARS-COV-2 IgG/IgM RDT in 31 PCR positive and 30 prospective negative samples. We compare the performance of the RDT in venous whole blood and separated plasma in a randomized blinded study and demonstrate that the tests show a significantly higher positivity agreement with PCR when plasma is used as the sample input. We also illustrate that H.E.R.M.E.S plasma is an advantageous sample type for use in serology testing as it widens the specific IgG detection window to within 2 weeks of infection. To further establish the validity of the study, we asked 10 previously untrained users to participate in a survey to assess the outcomes of the tests who also noted a similar increase in positivity agreement between the two sample types. The increase in performance as noted by trained professionals and untrained users with H.E.R.M.E.S plasma suggests that it is possible to avoid the issue of direct testing in blood altogether which could enhance the legitimacy of the use case for SARS-COV-2 serology RDTs in decentralized testing environments and facilitate the implementation of mass screening efforts to help mitigate the ongoing pandemic.

## Materials and Methods

### Sample Preparation

A total of 61 samples were used in this study (31 PCR positive and 30 presumed negative samples). PCR positive samples were acquired from two separate commercial collection services: CantorBio (San Diego, CA) and AMSBio (Cambridge, MA). Of the 31 PCR positive samples, 16 were collected with additional inclusion criteria of “Patients who tested PCR positive within 14 days prior to collection”. The 30 negative samples were acquired from Innovative Research (Novi, MI) and were further split into frozen plasma samples acquired prior to the pandemic and fresh whole blood samples collected prospectively from donors who had not yet tested PCR positive. For the purposes of equivalency, the frozen negative samples were mixed with pooled washed red blood cells (Innovative Research, Novi, MI) to mimic the consistency of a blood sample with a 50% hematocrit. Once the samples were received from their respective vendors, they were anonymized and assigned a random 6-digit alphanumeric code to facilitate a blinded study.

### Lateral Flow Rapid Diagnostic Tests

COVID 19 Rapid IgG/IgM tests (Healgen Scientific, San Diego, USA) were acquired from Confirm Biosciences (San Diego, CA). We compared the performance of the test in two different sample types: whole blood and plasma separated by the H.E.R.M.E.S platform. Briefly, the protocol for the whole blood test involved dispensing 10μL of sample into the input portion of the test strip. This was immediately followed by the application of 2 drops of buffer provided by the manufacturer. For the separated plasma samples, 20μL of whole blood was dispensed into a PCR tube containing the magnetic bead separation assay and placed in a small benchtop device for separation as outlined in Vemulapati et al (ref). After the plasma was separated, a capillary pipette was used to extract 5μl of plasma which was dispensed onto the sample input portion of the test. The test was then initiated by dispensing 2 drops of buffer. For either sample type, the test was allowed to run for 10 minutes before a positive or negative result was noted. Individual IgG and IgM responses were recorded based on the presence of red bands in the IgG and IgM “Test” line. Positive agreement with PCR was recorded if a red band was observed in either the IgG or IgM section of the test. A faint test line was also considered as a positive sample. Results were recorded by a trained laboratory professional. After the results of each test were recorded, the test strip was imaged by a rudimentary smartphone imaging setup using an Iphone 7 (Apple Inc, Cupertino CA) for use in the untrained user survey.

### Untrained User Survey

The smartphone images collected during testing were cataloged and input into a survey (Google Forms) to use in a short study to gather blinded performance data from previously untrained users. A total of 122 images (61 images for each sample type) were used for the evaluation. 10 participants were recruited using Amazon Mechanical Turk to participate in the study. The participants were required to have a high school degree at the very minimum in order to meet the suggested education requirement for laboratory personnel in a Certificate for Laboratory Improvement Amendment (CLIA) approved lab. There was no explicit mention that the images were collected from SARS-COV-2 serology tests.

To start the survey, participants were provided with brief instructions and background information necessary to classify the tests as positive, negative or invalid. The participants were then asked to complete a six-question training survey to help familiarize themselves with the selection criteria. Each question in the survey contained an image of a rapid test with either a sample positive for IgG alone, positive for IgM alone, positive for both IgG and IgM and negative for both IgG and IgM. For each training question, participants were asked to determine if the test was ‘Positive’, ‘Negative’ or ‘Indeterminate’. If a participant answered a question wrong in the training survey, they were informed of their error and given another chance to get the question right. The training survey was primarily designed to ensure that the participants had an accurate understanding of the classification criteria and was not used to judge results from the full survey. Once the training survey was completed, participants were advanced to the main section of the survey which included evaluating the full set of 122 images. These images were further divided into 6 sections in order for participants to accurately track their own progress. The order that images were displayed were also randomized for each participant.

## Results

### Comparison of performance in whole blood and H.E.R.M.E.S plasma

Due to the lack of a commercially available gold standard serology test for SARS-COV-2, we used a Positive Percentage Agreement (PPA) and Negative Percentage agreement (NPA) against PCR to evaluate diagnostic accuracy of the antibody test. PPA/NPA for the whole blood and H.E.R.M.E.S can be seen in Table 1 and Table 2. For the 31 positive samples, we note an increase in PPA from 83.9% in whole blood to 96.7% in separated plasma. The difference in PPA is further highlighted in the set of samples collected from patients within the first 14 days of PCR positive diagnosis (68.8% in whole blood and 93.8% in H.E.R.M.E.S plasma). The difference in PPA can be largely attributed to the false negatives from whole blood samples collected from patients with early stage infections, an example of which is shown in Figure 1A. The test correctly identified IgG in all 15 blood samples that were collected from patients within 1-2 months of infection. We also observed a significant increase in the IgM detection rate from 64.5% in whole blood to 80.6% in H.E.R.M.E.S plasma. The NPA remained at 100% for both sample types.

**Table 1:**
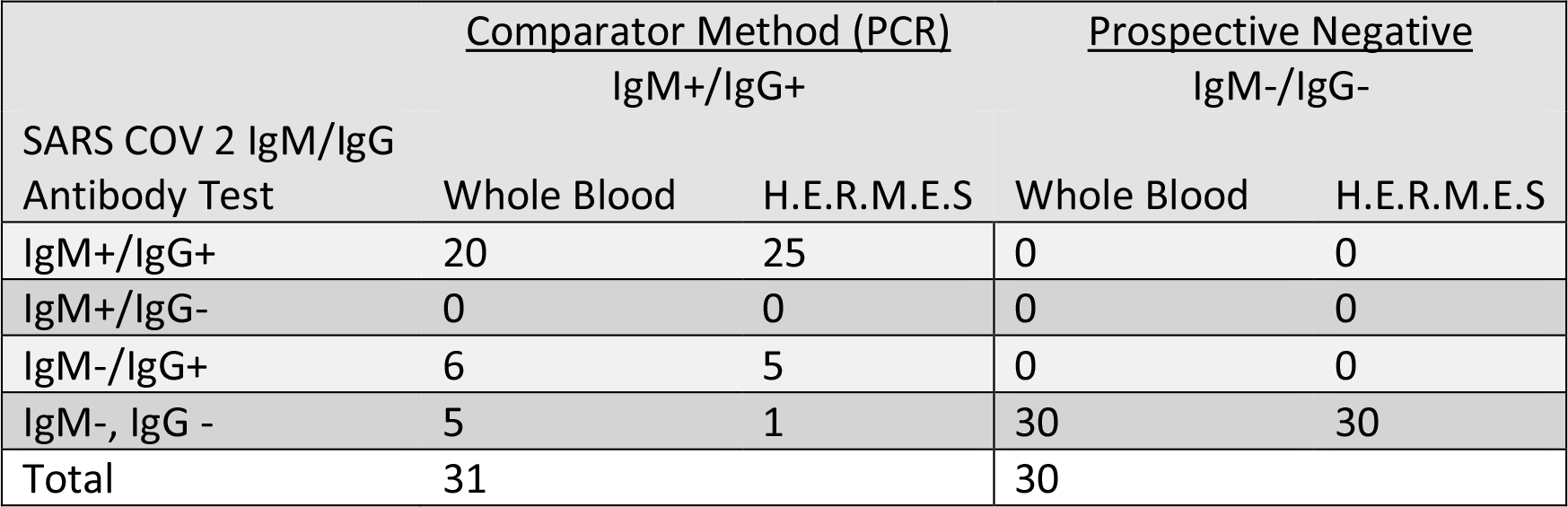
Summary of results for comparison between Whole blood and H.E.R.M.E.S plasma

**Table 2:** Comparison of PPA and NPA between Whole Blood and H.E.R.M.E.S. % represents agreement with PCR in the case of positives. C.I. represents the 95% Confidence interval.

|  | Whole Blood |  | H.E.R.M.E.S |  |
| --- | --- | --- | --- | --- |
|  | % | C.I. | % | C.I. |
| PPA | 83.9% (26/31) | (70.7%-97%) | 96.7% (30/31) | (90.5%-100%) |
| PPA<br>(early infections) | 68.8% (11/16) | (45.2%-92.2%) | 93.8% (15/16) | (81.5%-100%) |
| NPA | 100% (30/30) |  | 100% (30/30) |  |

**Figure 1:**
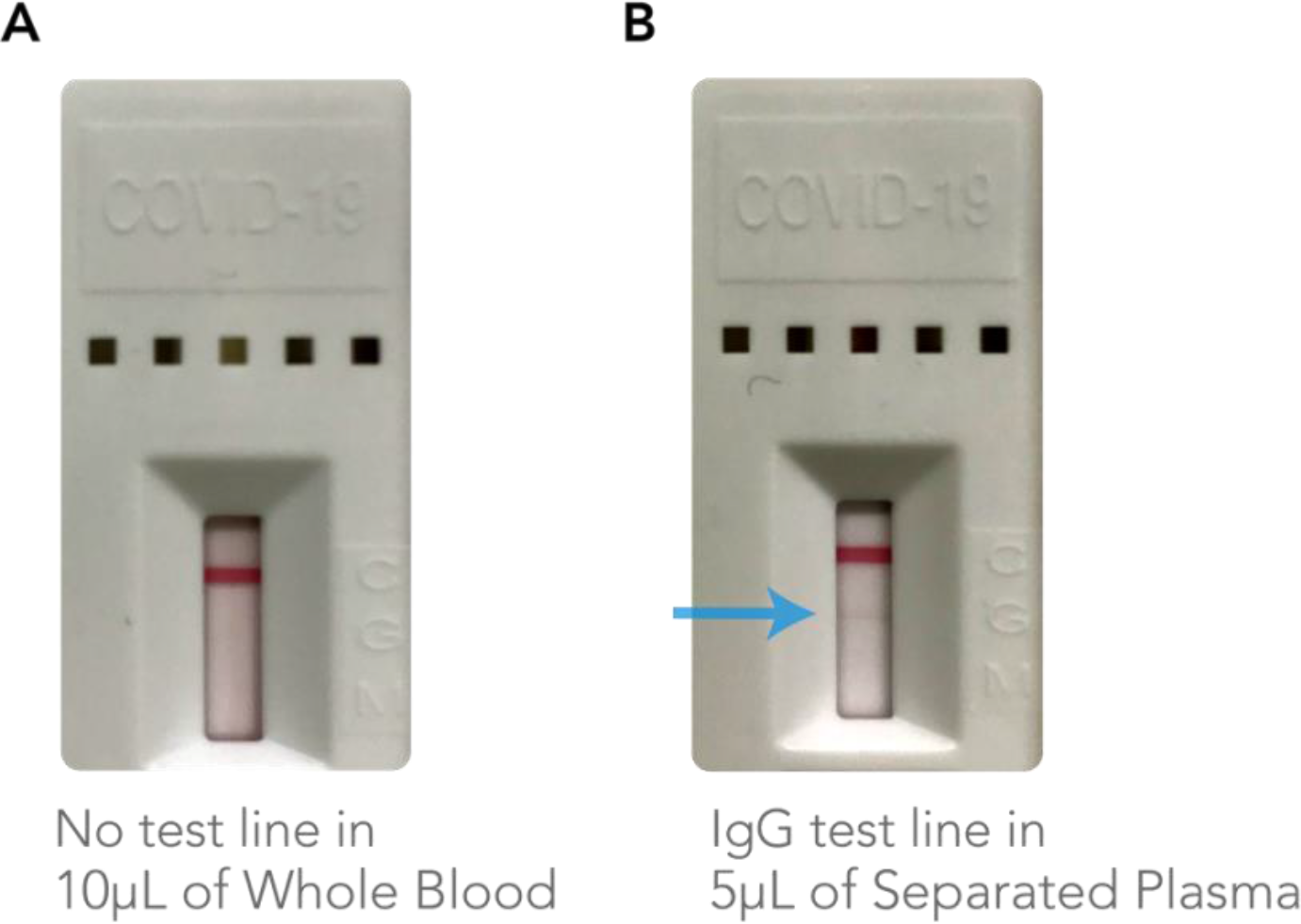
Example of an incorrectly assessed False Negative test in whole blood. A) Test result from using whole blood and B) test result using H.E.R.M.E.S plasma from the same sample. A faint IgG response is noted by the arrow.

### Untrained User Evaluation

Table 3 shows the average PPA and NPA as determined by the 10 previously untrained users. We note a slightly diminished increase in total PPA from whole blood to H.E.R.M.E.S (80.7% to 89.4%) and a similar increase in PPA for the early infections sample subset (62.6% to 93.1%). The NPA for both sample types were comparable (98.3% vs 99.7%). The Fleiss’ kappa for inter rater reliability^27^ was evaluated to be significantly greater than 0.81 in both cases demonstrating near perfect agreement between the evaluators across both sample types.

**Table 3:**
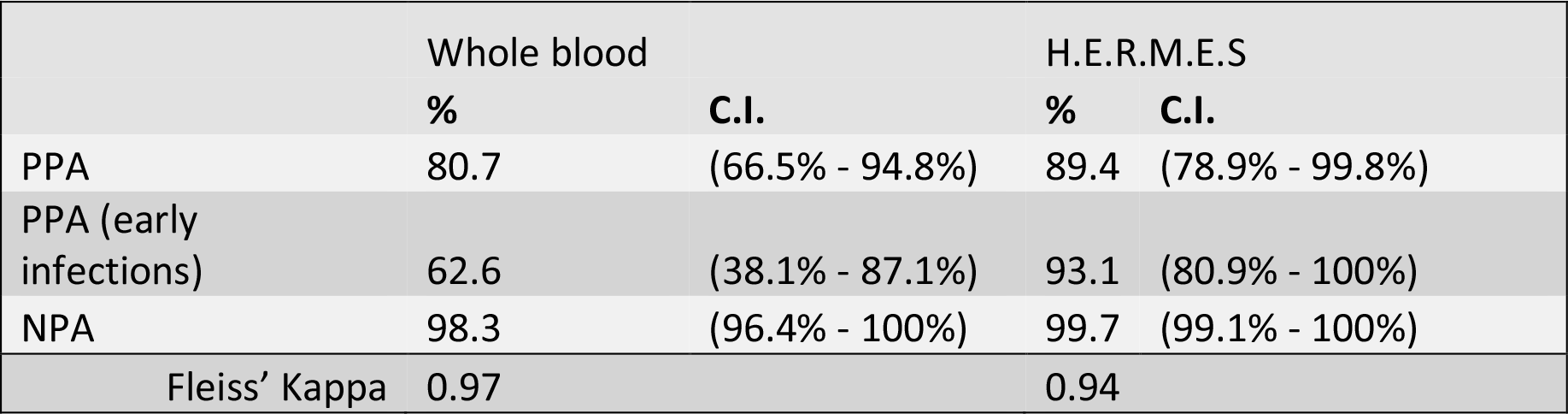
Comparison of PPA/NPA as determined by 10 untrained users. Fleiss’ Kappa for interrater reliability is shown in the last row.

|  | Whole blood |  | H.E.R.M.E.S |  |
| --- | --- | --- | --- | --- |
|  | % | C.I. | % | C.I. |
| PPA | 80.7 | (66.5% - 94.8%) | 89.4 | (78.9% - 99.8%) |
| PPA (early<br>infections) | 62.6 | (38.1% - 87.1%) | 93.1 | (80.9% - 100%) |
| NPA | 98.3 | (96.4% - 100%) | 99.7 | (99.1% - 100%) |
| Fleiss' Kappa | 0.97 |  | 0.94 |  |

## Discussion

Rapid serological screening at the point of care can enable local governments, communities and workplaces to effectively monitor and track the spread of SARS-COV-2 in their respective populations^28,29^. For successful implementation, it is critical that serology RDTs exhibit a high diagnostic accuracy (>95%) in venous or fingerstick blood, as they are likely the only accessible sample type at the point-of-care. In this work, we demonstrate the validity of a portable plasma separation system as an alternative approach to improve the performance of SARS-COV-2 serology RDTs for use at the point-of-care. By enabling plasma separation in decentralized testing environments, the H.E.R.M.E.S platform bypasses the need for arduous optimization of existing assays for use in blood. We offer evidence to suggest that plasma shows a significant improvement in positivity agreement against PCR when compared to whole blood and further demonstrate that the use of plasma can enhance the detection window for specific IgG to within 2 weeks of infection. We also demonstrate that the difference in observed performance is similar when the tests are evaluated by a laboratory professional and an untrained user, further validating the use case for implementing H.E.R.M.E.S with RDTs in decentralized testing environments. The ability to facilitate earlier detection and the capability to make plasma accessible at the point-of-care make a compelling case for the incorporation of the H.E.R.M.E.S plasma separation system for any widescale serology testing pursuit.

A higher positivity agreement rate with PCR would likely translate to a higher sensitivity for these tests in clinical settings. A highly sensitive serology RDT for SARS-COV-2 could be impactful in a variety of clinical situations. In addition to detecting a higher number of infections in patients who are within the first few days of infection, an enhanced sensitivity would also help inform infection history in those individuals who were infected during the onset of the pandemic. With an expectation of lowered levels of specific antibody in those who are several months removed from exposure to SARS-COV-2, a highly sensitive test could help accurately diagnose past infection and prevent more false negatives. Additionally, a point-of-care serology test could be extremely beneficial when an effective vaccine has been verified and deployed across large masses of the population. The ability to reliably verify antibody presence at the point-of-care could help state, county and city governments track the progress of their respective vaccination efforts. In turn, the means to quickly verify successful vaccination could lead to more robust pathways to help workplaces and business gradually return to optimal operating conditions.

The enhanced performance of RDTs in separated plasma can be largely attributed to the absence of red blood cells. In addition to interfering with the underlying immunochemistry, red cells make visual readout based on colorimetric bands difficult due to the increased background and noise due to the pigment of the cells. This also makes it harder to visually identify faint signal responses in samples that might have relatively low level of analyte. This is further exemplified in the higher IgM positivity rate in plasma over whole blood as noted in the results of the study. While faint lines can be difficult to recognize, they can be used to trigger a follow-up confirmatory test, a common practice employed in decentralized testing environments. The faint response could likely be solved by using a test from a different manufacturer or dispensing 30% more sample volume than recommended. We also expect that an automated imaging system could be a viable alternative to avoid the issue of human error altogether. In this study, all faint responses were classified as positive which could help explain the diminished increase in PPA determined by the untrained users in comparison to the laboratory professional.

The overall performance of the test in whole blood as determined in this study is much lower than described by the manufacturer and the National Cancer Institute’s independent evaluation in serum samples^30^. The difference in performance is more noticeable in the case of IgM. The diminished sensitivity in detection of specific IgM is expected in whole blood due to a combination of lower overall performance and the smaller amount of IgM generated by the body during an immune response as compared to IgG ^31^. However, we also suspect that the lower performance in IgM is a function of the design of multiplexed lateral flow assays; the architecture of the test strip tends to favor the detection of the analyte that is captured further away from the point of sample input due to the dependence of reaction time on flow rate^32^. As such, we expect the IgM performance could be improved with the design of a standalone IgM test or by employing a different test strip architecture. Finetuning the IgM responses could play a bigger role in enabling earlier detection, particularly when used in combination with a PCR test^33^.

In this work, we demonstrate the compatibility of the H.E.R.M.E.S platform with a single commercially available EUA approved SARS-COV-2 serology rapid test and we expect the performance difference to be similar in other serology tests that are compatible with plasma and whole blood. The difference in performance between blood and plasma will likely also depend on the recommended protocol outlined by the manufacturers in the “Instruction for Use” (IFU) documents. Specifically, the recommend sample volumes will dictate the difference in performance of the test as immunochemical tests are sensitive to changes in total analyte^34^. While some manufacturers suggest a specific amount for a given sample type (typically the volume of recommended plasma is half of the amount of blood), others recommend the use of the same amount of volume regardless of sample type. In the case of the latter, we expect the differences in performance between whole blood and plasma to be even greater than demonstrate here due to the higher total amount of analyte in the plasma sample.

The number of positive samples we chose to evaluate in this study is in line with the number of positives expected to be tested by the FDA as outlined in their EUA requirements. Although the EUA recommends testing a total of 75 negative samples for each new antibody test, we focused our efforts in this study to evaluate the differences in positive percentage agreement that would arise from a difference in sample type. Next steps will involve further validation for use at the point of care by conducting a fingerstick study with volunteers in a point-of-care clinic of a similar size to evaluate the applicability and of our device in settings outside of the lab. Future studies will also include evaluation of the test from samples with known underlying conditions that are known to cross react with the COVID test (e.g., H.I.V).

## Conclusion

We demonstrate here the clear value that H.E.R.M.E.S plasma presents as a sample type for use with existing SARS-COV-2 serology testing RDTs as opposed to whole blood. Incorporation of the separation platform into existing testing protocols can immediately enhance the legitimacy of the use case for rapid serology testing and circumvent the arduous need to demonstrate equivalent performance in blood. We expect the H.E.R.M.E.S platform to play an integral role in any large-scale testing efforts that will require the deployment of serology RDTs to communities across the country.

## Data Availability

Detailed data for this work can be requested by emailing

## Acknowledgments

SV would like to thank Serhat Pala at Confirm Biosciences for providing the Healgen antibody tests for the study.

